# Genome-wide association study of autopsy-confirmed Multiple System Atrophy identifies common variants near *ZIC1* and *ZIC4*

**DOI:** 10.1101/2021.11.11.21265915

**Authors:** Franziska Hopfner, Anja K. Tietz, Viktoria Ruf, Owen A. Ross, Shunsuke Koga, Dennis Dickson, Adriano Aguzzi, Johannes Attems, Thomas Beach, Allison Beller, Vivianna van Deerlin, Paula Desplats, Günther Deuschl, Charles Duyckaerts, David Ellinghaus, Valentin Evsyukov, Margaret Ellen Flanagan, Andre Franke, Matthew P. Frosch, Marla Gearing, Ellen Gelpi, Bernardino Ghetti, Jonathan D. Glass, Lea T. Grinberg, Glenda Halliday, Ingo Helbig, Matthias Höllerhage, Inge Huitinga, David John Irwin, Dirk C. Keene, Gabor G. Kovacs, Edward B. Lee, Johannes Levin, Maria J. Martí, Ian Mackenzie, Ian McKeith, Catriona Mclean, Brit Mollenhauer, Manuela Neumann, Kathy L. Newell, Alex Pantelyat, Manuela Pendziwiat, Annette Peters, Laura Molina Porcel, Alberto Rabano, Radoslav Matěj, Alex Rajput, Ali Rajput, Regina Reimann, William K. Scott, William Seeley, Sashika Selvackadunco, Tanya Simuni, Christine Stadelmann, Per Svenningsson, Alan Thomas, Claudia Trenkwalder, Claire Troakes, John Q. Trojanowski, Charles L. White, Tao Xie, Teresa Ximelis, Justo Yebenes, the Alzheimer’s Disease Genetics Consortium, Ulrich Müller, Gerard D. Schellenberg, Jochen Herms, Gregor Kuhlenbäumer, Günter Höglinger

## Abstract

Multiple System Atrophy is a rare neurodegenerative disease with alpha-synuclein aggregation in glial cytoplasmic inclusions and either predominant olivopontocerebellar atrophy or striatonigral degeneration, leading to dysautonomia, parkinsonism, and cerebellar ataxia. One prior genome-wide association study in mainly clinically diagnosed patients with Multiple System Atrophy failed to identify genetic variants predisposing for the disease. Since the clinical diagnosis of Multiple System Atrophy yields a high rate of misdiagnosis when compared to the neuropathological gold standard, we studied common genetic variation in only autopsy-confirmed cases (N = 731) and controls (N = 2,898).

The most strongly disease-associated markers were rs16859966 on chromosome 3 (P = 8.6 × 10^−7^, odds ratio (OR) = 1.58, [95% confidence interval (CI) = 1.32-1.89]), rs7013955 on chromosome 8 (P = 3.7 × 10^−6^, OR = 1.8 [1.40-2.31]), and rs116607983 on chromosome 4 (P = 4.0 × 10^−6^, OR = 2.93 [1.86-4.63]), all of which were supported by at least one additional genotyped and several imputed single nucleotide polymorphisms with P-values below 5 × 10^−5^. The genes closest to the chromosome 3 locus are *ZIC1* and *ZIC4* encoding the zinc finger proteins of cerebellum 1 and 4 (ZIC1 and ZIC4).

Since mutations of *ZIC1* and *ZIC4* and paraneoplastic autoantibodies directed against ZIC4 are associated with severe cerebellar dysfunction, we conducted immunohistochemical analyses in brain tissue of the frontal cortex and the cerebellum from 24 Multiple System Atrophy patients. Strong immunohistochemical expression of ZIC4 was detected in a subset of neurons of the dentate nucleus in all healthy controls and in patients with striatonigral degeneration, whereas ZIC4 positive neurons were significantly reduced in patients with olivopontocerebellar atrophy.

These findings point to a potential ZIC4-mediated vulnerability of neurons in Multiple System Atrophy.

## Introduction

Multiple System Atrophy (MSA) is a rapidly progressive rare neurodegenerative disease presenting with variable combinations of dysautonomia, parkinsonism and cerebellar ataxia[10]. Two forms of MSA can be clinically distinguished, characterized by either predominant parkinsonism (MSA-P) or predominant cerebellar symptoms (MSA-C)[12]. Its estimated prevalence is 3.4 to 4.9 cases per 100,000 individuals in the general population, and 7.8 cases per 100,000 in persons older than 40 years[32]. The mean survival time from disease onset is 6 to 10 years [16, 41]. Currently, only limited symptomatic treatments and no disease-modifying therapies are available[28].

The typical symptoms of MSA are caused by the progressive degeneration of neurons in different brain regions, particularly in the substantia nigra, striatum, inferior olivary nucleus, pons and cerebellum, but also other parts of the central nervous systems, emphasizing the multisystem character of MSA [12, 40]. The histological hallmarks in brains of MSA patients are glial cytoplasmic inclusions (GCIs, Papp-Lantos bodies) in oligodendrocytes containing aggregated and misfolded alpha-synuclein[14]. Neuropathologically, two subtypes can be distinguished, one with predominant olivopontocerebellar atrophy (OPCA), the other with mainly striatonigral degeneration (SND)[15, 26]. In addition, a mixed phenotype displaying features of both OPCA and SND is found in the brains of some patients[15, 26].

The pathogenesis of MSA is unclear. MSA is considered as a sporadic disease[11]. Epidemiological studies have investigated the influence of environmental factors in MSA, including exposure to farming-related factors (pesticides, solvents, mycotoxins, dust, fuels, oils, fertilizers, animals) and certain lifestyles (consumption of well water, rural living, diet and physical activity)[8, 23, 39]. Apart from a marginal effect of pesticides, no other environmental factors have been convincingly associated with an increased risk of developing MSA[8, 23, 39].

Hypothesis-driven candidate gene studies have been inconsistent with respect to variants that might be associated with MSA. Associations of MSA with the genes *COQ2, SNCA, MAPT*, and *PRNP* have been discussed[7, 20, 21, 29, 31, 33]. One prior genome-wide association study (GWAS) did not identify hits of statistical significance at a genome-wide level, despite the analysis of 918 cases and controls[30]. This GWAS had mainly included clinically diagnosed MSA cases. It needs to be stressed that clinical diagnosis is frequently not accurate in MSA. For example, a recent clinico-pathological study demonstrated a false positive diagnosis at autopsy in 38% of clinically diagnosed MSA patients[17].

In order to avoid inclusion of misdiagnosed patients in the GWAS described here, we only included autopsy-confirmed cases and appropriate ethnicity-matched controls.

## Methods

### Patient recruitment

Ethical approval had been obtained from all responsible ethics committees. All participants had given written consent.

Neuropathologists at each recruitment site (Supplementary material, Supplementary Table S1) based the definite neuropathological diagnosis of MSA on histopathological criteria taking into account GCIs immunoreactive for alpha-synuclein in characteristic anatomical distribution as defining feature of MSA[37]. Age, gender, disease history (including disease onset and duration) and neuropathological findings were recorded in standardized manner for all cases.

Controls were ethnically matched to cases and either derived from biobanks KORA-gen[42] or popGen[24] (Europe sites) or from North American site (Alzheimer’s Disease Genetics Consortium)[19]. The Alzheimer’s Disease Genetics Consortium assembled and genotyped DNA from subjects enrolled in the 29 NIA-Alzheimer’s Disease Centers located across the USA. For this study, the ADGC provided a subset of mostly clinical, cognitively normal controls. Patients and controls were of North-Western European and African American ancestry.

### DNA extraction

We isolated DNA from 30 mg frozen cerebellar cortex using QIAamp DNA Mini Kit (Qiagen, Venlo, Netherlands). DNA extraction was performed at German Center for Neurodegenerative Diseases (DZNE), Munich Germany. DNA was stored at -80 °C until use. DNA concentration was measured using a NanoDrop Spectrophotometer. DNA quality was determined by gel electrophoresis.

### Genotyping

All samples were genotyped on Infinium Global Screening Arrays (GSA, Illumina, San Diego, USA). The cases were genotyped at the Institute of Clinical Molecular Biology, Kiel University, Germany. The samples were genotyped in one batch on array version 2.0 for cases and version 1.0 for controls. Genotypes were called using Illumina Genome studio according to the manufacturers instructions using in-house cluster-files.

### Quality control and imputation

We used PLINK (v. 1.9) [1] and R (v. 3.6.3)[35] for all analyses. Only variants successfully genotyped in both the patient and the control populations were included in the subsequent analyses. Variants with multi-character allele codes, insertions, deletions, duplicated markers and all A/T and G/C variants were excluded. We excluded all samples discordant between reported and genotypic sex. Missing sex was imputed and samples with ambiguously imputed sex were discarded. After a first step of filtering out samples and variants with call rate of less than 85%, we excluded variants with an individual call rate of less than 98% in a second filtering step. Next, we removed variants with a minor allele frequency (MAF) below 0.01, significant deviation from Hardy-Weinberg equilibrium (HWE; P < 1 × 10^−6^) in controls or informative missingness (P < 1 × 10^−5^). Subsequently, we excluded individuals with a variant call rate of less than 98% or an outlying heterozygosity rate (mean ± 3 standard deviations). We used a pruned dataset containing only markers in low linkage-disequilibrium (LD) regions (pairwise r^2^ < 0.2) to test for duplicated individuals and cryptic relatedness (Pihat > 0.125) using pairwise genome-wide estimates of the proportion of identity by descent. Of each detected sample pair we excluded the individual with a lower call rate. Ethnical outliers were identified by a principal component analysis (PCA) together with the publically available 1000 Genomes data with known ethnicities.[2] Since the study population has genetically a mainly European ancestry, as ascertained by the PCA, we determined a European center and excluded samples more than 1.5 times the maximal European Euclidean distance away from this center. After a first association analysis of genotyped single nucleotide polymorphisms (SNPs) only, we inspected visually the cluster plots of all variants with a P-value below 1 × 10^−5^ and discarded variants without adequate cluster separation.

Imputation was carried out on the quality assured dataset using the TOPMed Imputation Server, resulting in 271,284,138 available SNPs.[34] Variants were again filtered for MAF and deviation from HWE in controls with the same thresholds as before. Additionally, SNPs with an imputation quality score R^2^ less than 0.7 were excluded, leaving 8,131,900 variants for analyses. As final step of the quality control procedure, we employed the R package PCAmatchR to ethnically match cases to controls with a 1:4 ratio to overcome possible difficulties with population stratification, leading to 3240 individuals for the analyses.[6]

### Association analysis

We used logistic regression to test the additive genetic model of each marker for association with disease status. Following scree plot analysis, we incorporated the first two dimensions of the PCA and sex as covariates. We used a genome-wide significance threshold of P < 5 × 10^−8^ and the threshold of P < 5 × 10^−6^ for suggestive association. Conditional analyses including in turn each SNP with a suggestive association as additional covariate were conducted to identify adjacent independent signals. Furthermore, we tested for clumps of correlated SNPs, i.e. to assess how many independent loci had been associated and determined the number of variants supporting the lead SNPs at each locus, i.e. variants that have a P-value < 5 × 10^−5^, are in LD (r2 ≥ 0.4) and not further than 250 kilobases (kb) away from the respective SNP. Visualization of the results was carried out with R and LocusZoom [27]for regional plots. Variant positions in this manuscript are reported on human genome version 38 (GRCh38/hg38).

### Immunohistochemistry on MSA patients’ brain

Formalin-fixed and paraffin embedded (FFPE) tissue from MSA patients and controls without neurological or psychiatric diseases was obtained from the Neurobiobank Munich (NBM; Germany). All autopsy cases of the NBM were collected on the basis of an informed consent according to the guidelines of the ethics commission of the Ludwig-Maximilians-University Munich, Germany (# 345-13). MSA cases had been diagnosed according to established histopatological diagnostic criteria.[26, 37]

For ZIC4 immunohistochemistry, 5 µm thick sections of FFPE tissue of the frontal cortex and the cerebellar hemisphere including the dentate nucleus were prepared. After deparaffinization, heat-induced epitope retrieval was performed in Tris/EDTA, pH 9 at 95 °C for 30 minutes. For blocking of endogenous peroxidase and unspecific protein binding, the sections were incubated with 5% H_2_O_2_ in methanol for 20 minutes and I-Block reagent (Applied Biosystems, Waltham, MA, USA) for 15 minutes. Subsequently, ZIC4 primary antibody (rabbit, polyclonal, Merck/Sigma-Aldrich, Darmstadt, Germany) was applied over night at 4 °C at a dilution of 1:100. Signal detection was performed using the DCS *Chromo*Line DAB kit (DCS, Hamburg, Germany) according to the manufacturer’s instructions. Sections were counterstained for 1 minute with Mayer’s hemalum solution (Waldeck, Münster, Germany).

To determine the fractions of ZIC4 positive neurons of all neurons in the dentate nucleus, stained slides were scanned using a slide scanner (Axio Scan. Z1, Zeiss, Oberkochen, Germany) and visualized using the free ZEN lite software (v 3.3; Zeiss, Oberkochen, Germany). For statistical evaluation of the data, Student’s t-test was used, statistical significance was defined as P < 0.05.

## Results

### Patient sample

From the initial sample of 731 cases, 13 cases had to be excluded due to insufficient tissue quality. After thorough quality control and filtering, 648 cases and 2,592 controls covering 8,131,900 variants were included in the association analysis (Supplementary Fig. S1). The number of excluded samples and variants in each step of the quality control procedure is shown in Supplementary Tables S2 and S3.

### Genotyping

We performed logistic regression incorporating sex and determined the first two dimensions of PCA as covariates using the scree plot method. The genomic inflation factor of λ = 1.01 (unimputed λ = 1.01, Supplementary Fig. S2) indicates that no significant population stratification was present (Fig. 1a). We did not identify any disease-associated variants with a P-value below the genome-wide significance threshold of P < 5 × 10^−8^, but suggestive associations with P < 5 × 10^−6^ at ten different loci (Fig. 1b) with the leading SNP at each locus shown in Table 1. Conditional analyses including in turn any SNP with P < 5 × 10^−6^ excluded the presence of multiple independent signals at each locus. All variants with suggestive associations are listed in Supplementary Table S4. The most noteworthy hits were rs16859966 on chromosome 3 (P = 8.6 × 10^−7^, odds ratio (OR) = 1.58, [95% confidence interval (CI) = 1.32-1.89]), rs7013955 on chromosome 8 (P = 3.7 × 10^−6^, OR = 1.8 [1.40-2.31]) and rs116607983 on chromosome 4 (P = 4.0 × 10^−6^, OR = 2.93 [1.86-4.63]), which were supported by at least one additional genotyped as well as serveal imputed SNPs with P-values below 5 × 10^−5^ as discovered in the clumping analysis (Table 1). The genes closest to the chromosome 3 locus are the Long Intergenic Non-Protein Coding RNA 2032 (*LINC02032*) approximately 100 kb downstream and the zinc finger proteins of cerebellum 1 and 4 genes (*ZIC1, ZIC4*), located roughly 600 kb upstream (Fig. 1c). The top SNP rs7013955 on chromosome 8 maps to the lysyl oxidase-like 2 gene (*LOXL2*; Fig. 1d). The association signal around SNP rs116607983 on chromosome 4 is located in a region devoid of protein-coding genes approximately 2000 kb to either side (Figure 1e). A fourth locus on chromosome 5 (rs2279135) was also supported by multiple clumped SNPs, but all SNPs including the lead SNP were imputed (Table 1). Several variants clumped at the chromosome 5 locus were located in the *ARHGEF37* gene, coding for Rho Guanine Nucleotide Exchange Factor 37 (Fig. 1f). None of the identified SNPs is an expression quantitative trait locus (eQTL) in brain tissues according the Genotype Tissue Expression (GTEx) project.[9] At four of the six remaining loci with variants exhibiting suggestive associations at most two supporting SNPs were present, which were all imputed; in the other two loci no supporting SNPs could be found in the clumping analysis (Table 1, Supplementary Fig. S3). We did not investigate these loci further, because it is unlikely that they represent valid associations. No suggestive associations were detected with previously reported Parkinson’s disease associations from a meta-analysis of 17 datasets from Parkinson’s disease GWAS (Supplementary Table S5).[22]

**Table 1.** Top SNPs at each locus with P < 5 × 10^−6^. Results from association analysis with logistic regression including sex and the first two dimensions of principal component analysis (PCA) as covariates in 648 cases with MSA and 2,898 controls. Only the leading single nucleotide polymorphism (SNP) at each locus with a suggestive association between the disease status and a variant is reported. Supplementary Table S3 lists all suggestive associations. BP = base-pair coordinates according to human reference genome GRCh38, CHR = chromosome, dbSNP = database of single nucleotide polymorphisms, GT = genotyped, IM = imputed, MAF = minor allele frequency.

| CHR | dbSNP ID | BP | MAF |  | OR [95% CI] | P | IM/<br>GT | # SNPs in Clump |  |  |
| --- | --- | --- | --- | --- | --- | --- | --- | --- | --- | --- |
|  |  |  | Cases | Controls |  |  |  | Total | GT | IM |
| 10 | rs4933352 | 85280795 | 0.42 | 0.52 | 0.71 [0.62-0.80] | 9.7E-08 | IM | 2 | 0 | 2 |
| 16 | rs79418449 | 80515374 | 0.04 | 0.02 | 2.54 [1.77-3.63] | 3.7E-07 | GT | 1 | 1 | 0 |
| **3** | **rs16859966** | **147976678** | **0.17** | **0.12** | **1.58 [1.32-1.89]** | **8.6E-07** | **IM** | **45** | **24** | **21** |
| 5 | rs114019803 | 159559041 | 0.02 | 0.01 | 3.36 [2.03-5.56] | 2.3E-06 | IM | 3 | 0 | 3 |
| 4 | rs933953 | 31356173 | 0.25 | 0.32 | 0.71 [0.62-0.82] | 2.6E-06 | IM | 1 | 0 | 1 |
| 18 | rs116914137 | 30589500 | 0.05 | 0.02 | 2.17 [1.57-3.00] | 2.8E-06 | IM | 3 | 0 | 3 |
| **8** | **rs7013955** | **23343590** | **0.08** | **0.05** | **1.80 [1.40-2.31]** | **3.7E-06** | **IM** | **20** | **1** | **19** |
| **4** | **rs116607983** | **33372461** | **0.03** | **0.01** | **2.93 [1.86-4.63]** | **4.0E-06** | **IM** | **88** | **3** | **85** |
| 11 | rs141819348 | 47698235 | 0.05 | 0.03 | 2.10 [1.53-2.88] | 4.6E-06 | IM | 3 | 0 | 3 |
| **5** | **rs2279135** | **149637742** | **0.32** | **0.27** | **1.39 [1.21-1.60]** | **4.8E-06** | **IM** | **24** | **0** | **24** |

**Fig. 1.**
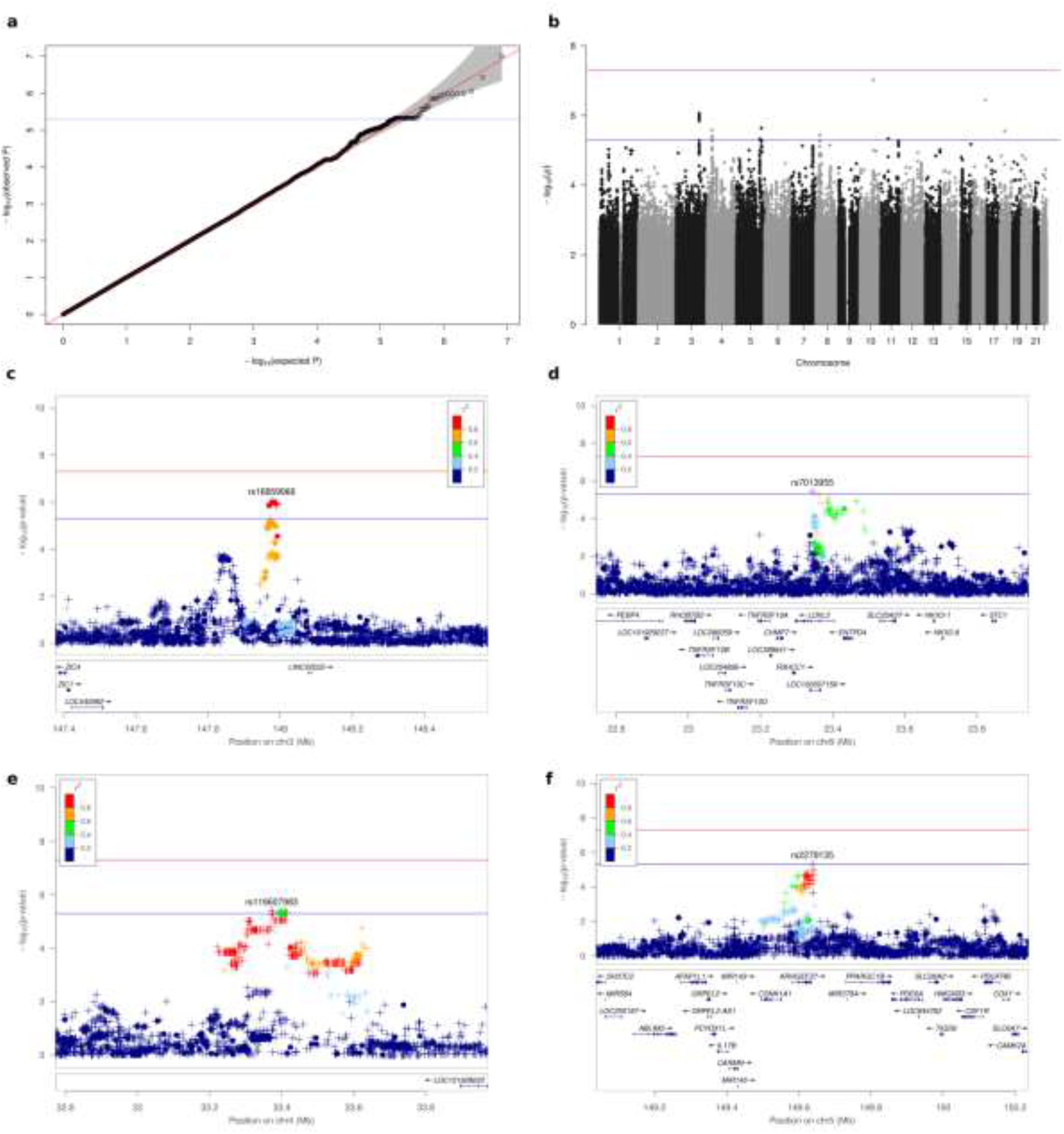
Association plots for MSA. **(a)** QQ (quantile-quantile) plot of association analysis for 8,109,760 variants. (**b)** Manhattan plot showing –log10 marker-wise P-values of the association analysis against their genomic base-pair position. The red and blue lines indicate the genome-wide significance threshold of 5 × 10^−8^ and threshold for suggestive associations of 5 × 10^−6^ respectively. (**c)** Regional plot for the association between MSA and variants on chromeome 3 in the genomic region from 147.4 to 148.6 Mb. A circle represents a genotyped and a plus symbol an imputed variant. The r^2^ metric displays the parwise LD between the leading and the resprective variant. The bottom part shows gene postions. **(d)** Regional plot for associations on chromosome 8 in the genomic resion from 22.7 to 23.9 Mb. **(e)** Regional plot for associations on chromsome 4 in the genomic region from 32.8 to 34.0 Mb. **(f)** Regional plot for associations on chromosome 5 in the genomic region from 149.0 to 150.2 Mb.

### ZIC4 immunohistochemistry on MSA patients’ brain

*ZIC4* and *ZIC1* are known to play a critical role in the embryonal development of the cerebellum. Heterozygous deletions comprising the *ZIC1* and *ZIC4* locus have been associated with the Dandy-Walker malformation, a rare congenital condition characterized by a hypoplastic cerebellar vermis and an enlarged fourth ventricle.[5, 13] In mice, deletions of *ZIC1* and *ZIC4* lead to a striking phenotype similar to the Dandy-Walker malformation with cerebellar hypoplasia and foliation defects.[5, 13] In addition, paraneoplastic autoantibodies against ZIC4 protein are linked to severe cerebellar dysfunction and degeneration.[3, 4]

Since cerebellar degeneration and corresponding symptoms are also a central hallmark of MSA, we decided to follow up on a potential role of ZIC4 in MSA patient brains by performing immunohistochemical stainings. For ZIC1 no primary antibody was appropriately sensitive and specific on human tissue in our hands. Thus, FFPE tissue of the frontal cortex and the cerebellum of MSA patients (N=10 SND, N=14 OPCA/mixed phenotype) and healthy controls (N=5) were stained with antibodies raised against ZIC4.

Nuclear and cytoplasmic staining of frontal cortex neurons was observed in all brains examined without differences between healthy controls and MSA patients (Fig. 2 a-c). In the cerebellar dentate nucleus, we found strong expression of ZIC4 in a subset of neurons in healthy controls as well as MSA patients with predominant SND (Fig. 2 d,e,g,h). In contrast, MSA patients with mixed subtype or OPCA showed reduced numbers of ZIC4-positive neurons, which were furthermore only weakly stained (Fig. 2 f, i). Quantification of the proportions of ZIC4-positive neurons among all dentate nucleus neurons revealed relatively constant proportions in healthy controls and patients with MSA-SND (33.2% ± 0.0 vs. 32.6% ±0.0), whereas in patients with MSA-OPCA or MSA-mixed phenotype we found significantly lower percentages of ZIC4-positive neurons (15.5% ± 0.1) (Fig. 2 j).

**Fig. 2.**
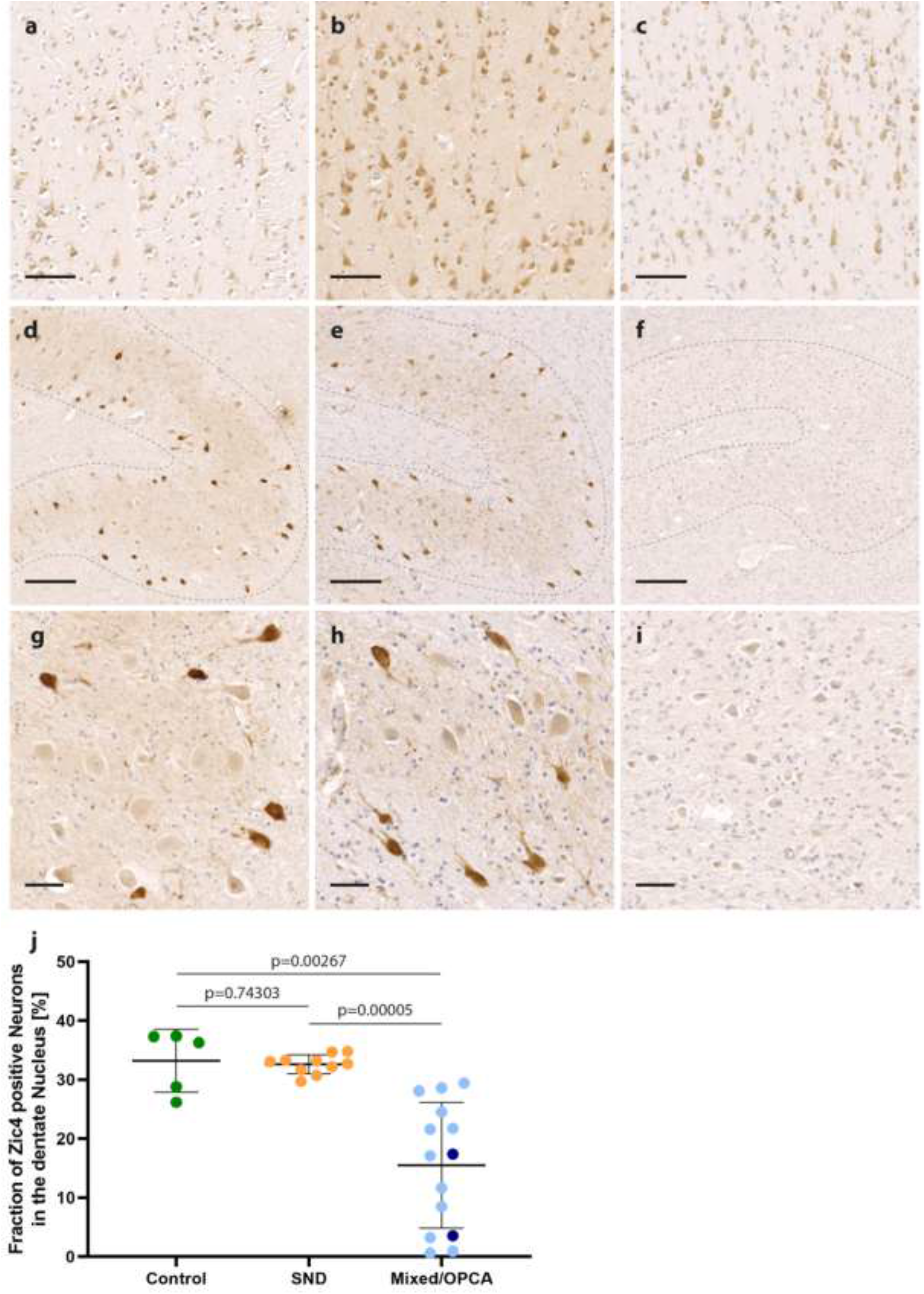
ZIC4 immunohistochemical staining of MSA patients’ and control brains. Representative ZIC4 immunohistochemical stainings of a control without neurodegenerative diseases **(a, d, g)** and two MSA patients with SND (striatonigral degeneration) **(b, e, h)** and a mixed subtype **(c, f, i)**, respectively. **(a-c)** Nuclear and cytoplasmic expression of ZIC4 was detected in a comparable manner in the frontal cortex of healthy controls and MSA patients **(d-i, higher magnification g-i)**. In the cerebellar dentate nucleus of healthy controls and patients with SND, a constant subset of neurons stained strongly positive for ZIC4, whereas in patients with OPCA (olivopontocerebellar atrophy) or mixed subtypes, only weak staining could be observed and the number of Zic4-positive neurons was clearly reduced. **(j)** Quantification of positive neurons in the dentate nucleus revealed a significant reduction of ZIC4-positive neurons in patients with either mixed subtype (light blue) or OPCA (dark blue) compared to SND or controls without neurodegenerative disease, while no difference was seen between patients with SND and healthy controls. Scale bars: a-c: 100 µm, d-f: 200 µm, g-i: 50 µm

## Discussion

As part of the study, brain banks were contacted worldwide and all available caucasian MSA brains were included. As in the prior GWAS with 918 predominantly clinically diagnosed MSA patients, our current GWAS of 648 autopsy-confirmed MSA patients, did not identify disease-associated common variants below the genome-wide significance threshold.

Since our prior GWAS of 219 patients with autopsy-confirmed corticobasal degeneration did identify significant disease-associated common variants, our current findings strongly suggest that the genetic contribution to disease risk is smaller in MSA.[18]

Nevertheless, our study reveals several suggestive associations at different loci, which may provide relevant hypotheses for follow-up investigations into the pathogenesis of MSA.

Specifically, we identified a variant on chromosome 3 (rs16859966, P = 8.6 × 10^−7^,, OR = 1.58 [1.32-1.89]) located upstream *ZIC1* and *ZIC4. ZIC1* and *ZIC4* are located in close genomic proximity to each other and encode transcription factors highly expressed in different brain areas[1, 38].

Proper function of these proteins is critical for the development of the CNS, particularly the cerebellum.[5] Although no effect of rs16859966 on *ZIC1* or *ZIC4* expression is recorded in the GTEx database, rare genetic variants or deletions in *ZIC1* or *ZIC4* result in congenital cerebellar defects.[5, 13, 25] A heterozygous deletion of *ZIC1* and *ZIC4* causes the Dandy-Walker malformation, a developmental disorder of the cerebellum.[13, 36] Moreover, paraneoplastic autoantibodies against ZIC4 induce cerebellar degeneration.[4] Due to the pronounced cerebellar degeneration in MSA, we followed-up on a possible role of ZIC4 in MSA.

While we could detect a relatively constant proportion of approximately one third ZIC4-positive neurons among all neurons in the cerebellar dentate nucleus in healthy controls and patients with MSA-SND, cases with MSA-OPCA or the mixed MSA phenotype showed significantly lower fractions of ZIC4-positive neurons. This finding suggests that ZIC4 may be involved in the neurodegeneration in MSA. The involvement of *ZIC4* mutations in the Dandy-Walker cerebellar malformation and the paraneoplastic ZIC4-autoantibody-associated cerebellar degeneration could suggest a pathomechanism in MSA, by which altered *ZIC4* expression could increase neuronal vulnerability. Further analyses of a potential functional interaction of alpha-synuclein and ZIC4 are currently ongoing.

We would like to strongly encourage independent replication studies to confirm or refute the hypotheses provided by our study.

## Data Availability

All data produced in the present work are contained in the manuscript

## Acknowledgements

We would like to thank all those who contributed towards our research, particularly the patients and families who donated brain tissue - without their donation this study would not have been possible. We thank Vanessa Boll and Lena Jaschkowitz for excellent technical assistance.

The study received support from the Deutsche Parkinson Gesellschaft, the Else-Kröner-Fresenius-Stiftung, the CurePSP foundation, the Alzheimer’s Disease Genetics Consortium (ADGC) NIA grant U01AG032984, the National Alzheimer’s Coordinating Center (NACC) NIA grant U01 AG016976 and the Deutsche Forschungsgemeinschaft (DFG, German Research Foundation) under Germany’s Excellence Strategy within the framework of the Munich Cluster for Systems Neurology (EXC 2145 SyNergy – ID 390857198), the Hannover Cluster RESIST (EXC 2155 – ID 390874280) and the Kiel Cluster (EXC2167, Precision Medicine in Chronic Inflammation (PMI)), the DFG grant (HO2402/18-1), the VolkswagenStiftung (Niedersächsisches Vorab), the Petermax-Müller Foundation (Etiology and Therapy of Synucleinopathies and Tauopathies), and ERARE18-124 (MSA-omics) under the frame of E-Rare-3, the ERA-Net for Research on Rare Diseases.

Data for this study were prepared, archived, and distributed by the National Institute on Aging Alzheimer’s Disease Data Storage Site (NIAGADS) at the University of Pennsylvania (NIA grant U24-AG041689).

Samples from the National Cell Repository for Alzheimer’s Disease (NCRAD), which receives government support under a cooperative agreement grant (U24 AG21886) awarded by the National Institute on Aging (NIA), were used in this study. We thank contributors who collected samples used in this study, as well as patients and their families, whose help and participation made this work possible;

A part of the samples was collected at the Mayo Clinic. The Mayo Clinic is an American Parkinson Disease Association (APDA) Mayo Clinic Information and Referral Center, an APDA Center for Advanced Research, and is supported by a Lewy Body Dementia Center Without Walls U54NS110435 (to D.W.D. & O.A.R.). The brain bank was supported, in part, by the Mangurian Foundation Lewy Body Dementia Program at Mayo Clinic.

The London Neurodegenerative Diseases Brain Bank, King’s College London is supported by the MRC (Medical Research Council, UK) and the Brains for Dementia Research project (jointly funded by the Alzheimer’s Society and Alzheimer’s Research UK).

Data were contributed to this study by the Center on Alpha-synuclein Strains in Alzheimer Disease & Related Dementias at the University of Pennsylvania Perelman School of Medicine (U19 AG062418, Trojanowski JQ-PI) and the former Morris K. Udall Center at the University of Pennsylvania Perelman School of Medicine (P50 NS053488, Trojanowski JQ-PI), NIA P01-AG066597 and AG072979 (formerly AG010124).

## Supplementary Tables

**Supplementary Table S1:** Recruitment centers, brain bank sources.

| City, Country | Source | MSA cases |
| --- | --- | --- |
| Zürich, Switzerland | Institute of Neuropathology, University Hospital of Zurich, Zurich, Switzerland. | 1 |
| Göttingen, Germany | University Medical Center Goettingen, Department of Neurology and Paracelsus-Elena-Klinik, 34128 Kassel, Germany | 2 |
| San Francisco, USA | University of California, San Francisco, CA, USA. | 2 |
| Vancouver, BC, Canada | University of British Columbia, Department of Pathology and Laboratory Medicine | 2 |
| New York, USA | Mount Sinai NBTR | 3 |
| Atlanta, USA | Emory University, Department of Neurology & Pathology | 4 |
| Los Angeles, USA | The Human Brain and Spinal Fluid Resource Center | 4 |
| Stockholm, Sweden | Department of Neurology, Karolinska University Hospital, Stockholm, Sweden. | 4 |
| Vienna, Austria | Institute of Neurology, Medical University of Vienna | 6 |
| Newcastle upon Tyne, UK | Newcastle Brain Tissue Resource, Newcastle University, Campus for Ageing and Vitality, Newcastle upon Tyne, NE4 5PL, UK | 7 |
| Chicago, USA | University of Chicago, Department of Neurology | 8 |
| Indiana, USA | Indiana University School of Medicine | 8 |
| San Diego, USA | San Diego Shiley-Marcos AD Research Center, University of California | 8 |
| Tübingen, Germany | Department of Neuropathology, University Hospital of Tübingen, Tübingen, Germany | 8 |
| Madrid, Spain | Centro de Biología Molecular "Severo Ochoa", c/Nicolás Cabrera, 1, Universidad Autónoma de Madrid, Cantoblanco, Madrid, Spain | 10 |
| Seattle, USA | Department of Pathology, University of Washington, Seattle, WA, USA | 10 |
| Prague, Czech Republic | Department of Pathology and Molecular Medicine, Thomayer University Hospital, Prague | 12 |
| Sidney, Australia | Brain and Mind Centre, Sydney Medical School, The University of Sydney, Sydney, NSW, Australia. | 12 |
| Arizona, USA | Banner Sun Health Research Institute, | 13 |
| Parkville, Victoria, Australia | Australian Brain Bank Network, Howard Florey Laboratories, The Florey Institute of Neuroscience and Mental Health | 13 |
| Dallas, Texas, USA | Alzheimer's Disease Center, University of Texas Southwestern Medical Center, Dallas, Texas, USA | 15 |
| Saskatchewan, Canada | Saskatoon Health Region/University of Saskatchewan, Rosthern; and Movement Disorders | 17 |
| Paris, France | Raymond Escourolle Neuropathology Department. Groupe Hospitalier Pitié-Salpêtrière, Paris, France | 20 |
| London, UK | Imperial College London | 22 |
| Baltimore, USA | Johns Hopkins Medical Institution Brain Resource Center, MD, USA | 24 |
| London, UK | MRC London Neurodegenerative Diseases Brain Bank, Institute of Psychiatry, Psychology and Neuroscience, King's College | 26 |
| Munich, Germany | Neurobiobank Munich, Center for Neuropathology and Prion Research, Ludwig-Maximilians University | 29 |
| Boston, USA | Massachusetts General Hospital | 30 |
| Barcelona, Spain | Neurological Tissue Bank of the Biobanc-Hospital Clinic-IDIBAPS | 34 |
| Amsterdam, Netherlands | Alzheimer Center | 36 |
| Ann Arbor, USA | University of Michigan, Department of Pathology, University of Michigan Medical School, Ann Arbor, MI, USA | 37 |
| Miami, USA | UM Brain Endowment Bank, an NIH NeuroBioBank | 45 |
| Pennsylvania, USA | The Penn FTD Center - University of Pennsylvania, USA | 54 |
| Jacksonville, Florida, USA | Department of Neuroscience, Mayo Clinic, Jacksonville | 205 |
| **Total** |  | **731** |

**Supplementary Table S2:** Summary of per sample quality control. SD = standard deviation.

|  | Cases | Controls | Total |
| --- | --- | --- | --- |
| **Samples started with** | **731** | **2,898** | **3,629** |
|  | Number of excluded samples |  |  |
| Insufficient brain tissue quality | 13 | 0 | 13 |
| Duplicate Samples | 2 | 0 | 2 |
| Sample call rate < 85% | 18 | 0 | 18 |
| Sample Call rate < 98% | 2 | 3 | 5 |
| Heterozygosity Rate > Mean ± 3xSD | 25 | 20 | 45 |
| Cryptic relatedness > third degree relatives | 4 | 33 | 37 |
| Missing or discrepant sex | 2 | 9 | 11 |
| Ethnical Outlier | 17 | 0 | 17 |
| Control Matching | - | 241 | 241 |
| Total samples excluded | 68 | 306 | 374 |
| **Total samples analysed** | **648** | **2,592** | **3,240** |

**Supplementary Table S3:**
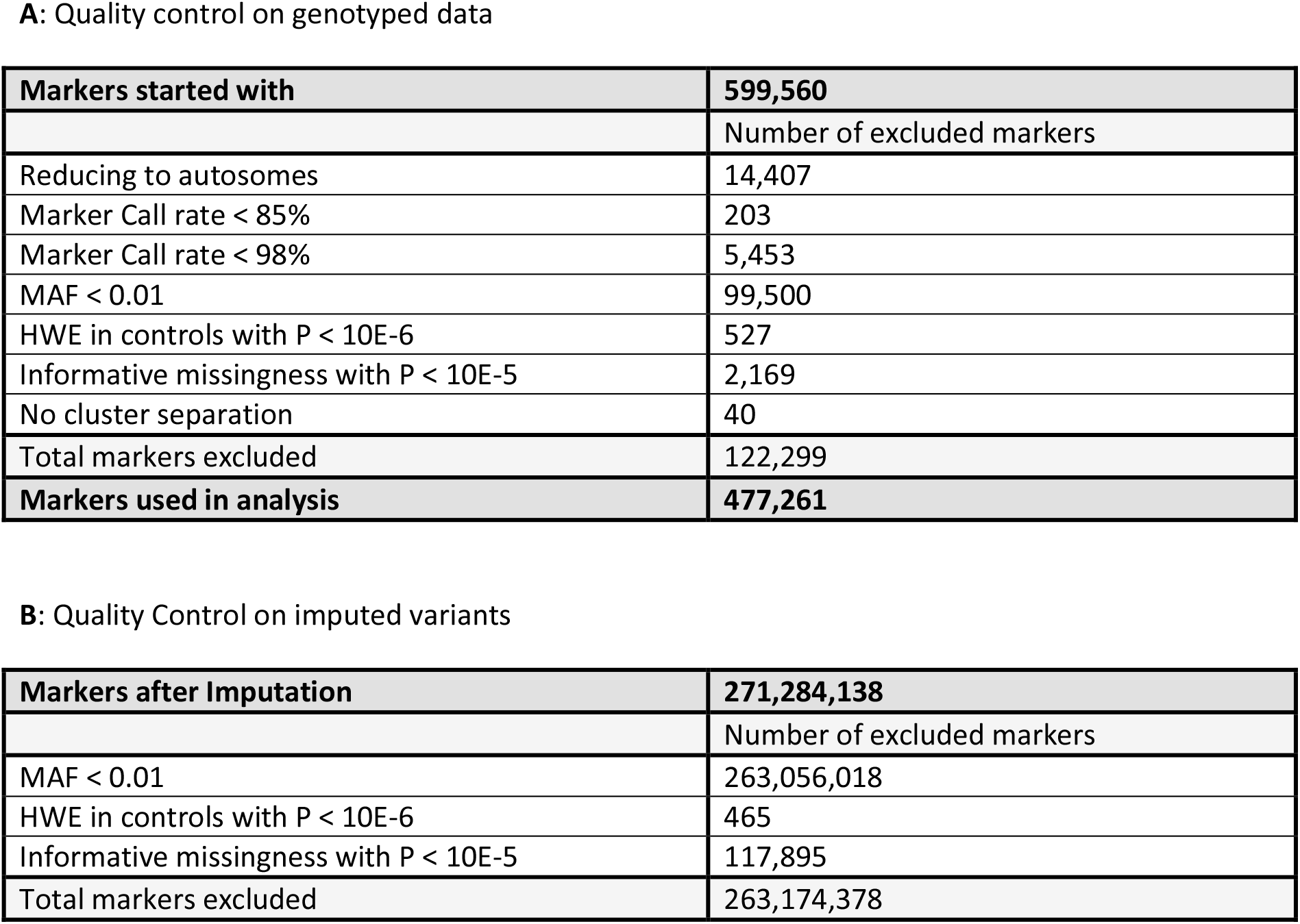
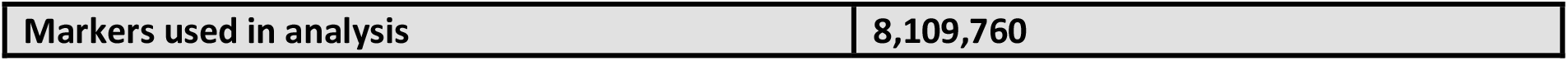
Summary of per marker quality control. MAF = minor allele frequency, HWE = Hardy-Weinberg equilibrium.

|  |  |
| --- | --- |
| **Markers started with** | **599,560** |
|  | Number of excluded markers |
| Reducing to autosomes | 14,407 |
| Marker Call rate < 85% | 203 |
| Marker Call rate < 98% | 5,453 |
| MAF < 0.01 | 99,500 |
| HWE in controls with $P < 10E-6$ | 527 |
| Informative missingness with $P < 10E-5$ | 2,169 |
| No cluster separation | 40 |
| Total markers excluded | 122,299 |
| **Markers used in analysis** | **477,261** |
| **B:** Quality Control on imputed variants |  |
| **Markers after Imputation** | **271,284,138** |
|  | Number of excluded markers |
| MAF < 0.01 | 263,056,018 |
| HWE in controls with $P < 10E-6$ | 465 |
| Informative missingness with $P < 10E-5$ | 117,895 |
| Total markers excluded | 263,174,378 |
| Markers used in analysis | 8,109,760 |

**Supplementary Table S4:**
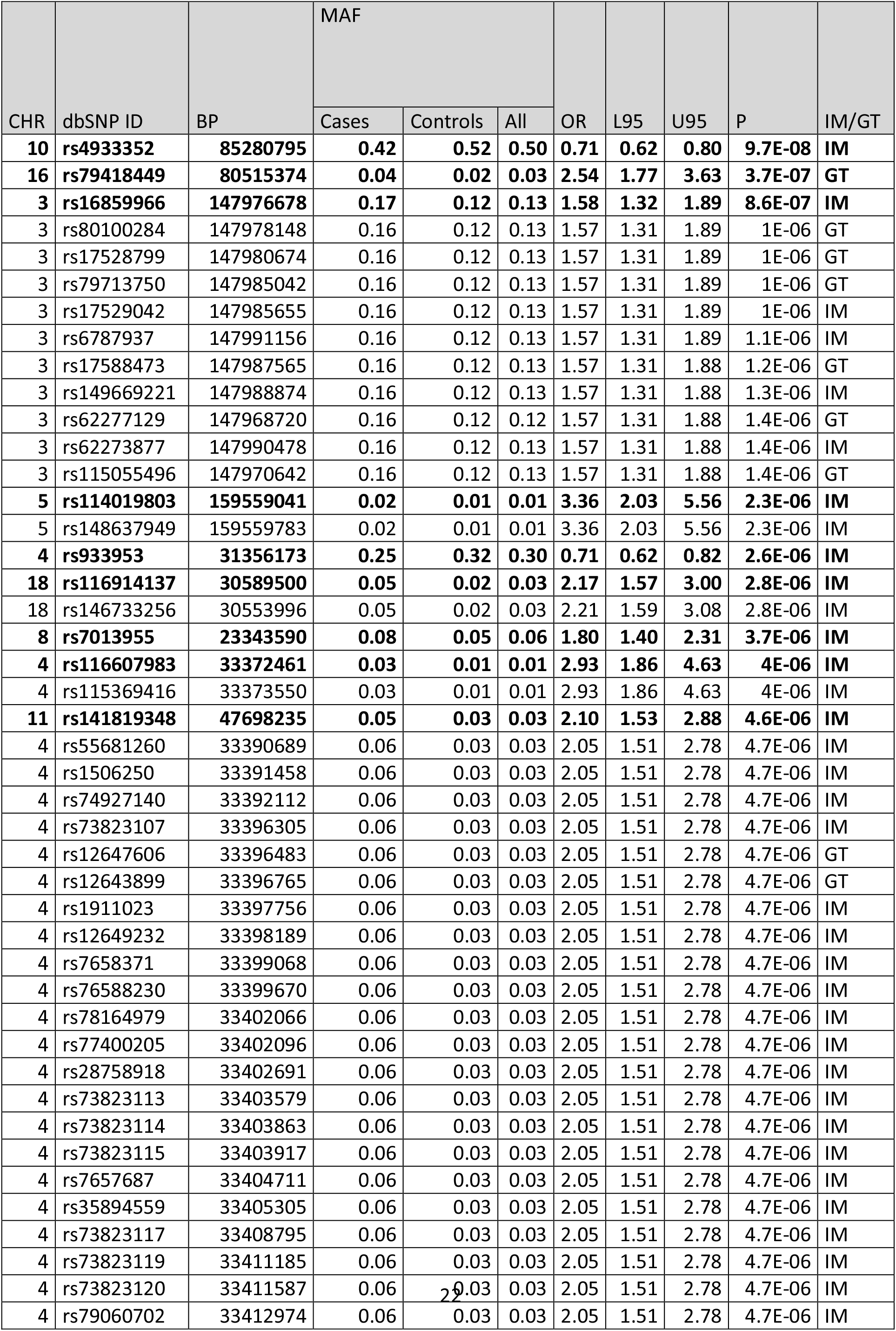
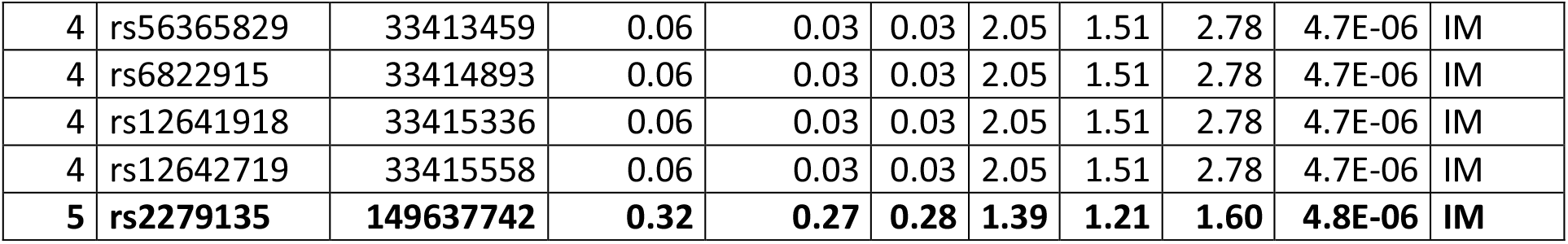
Identified associations with P < 5 × 10^−6^. Results from association analysis with logistic regression including sex and the first two dimensions of PCA as covariates in 648 cases with MSA and 2,898 controls. BP = base-pair coordinates according to human reference genome GRCh38, CHR = Chromosome, dbSNP = database of single nucleotide polymorphisms, GT = genotyped, IM = imputed, L95 = Lower bound of 95% confidence interval for odds ratio, MAF = minor allele frequency, U95= Upper bound of 95% confidence interval for odds ratio.

**Supplementary Table S5:** Top Parkinson’s disease SNPs according to the Parkinson’s disease meta-analysis of genome-wide association studies, Nalls et al. 2019. The SNP selection is based on results from the meta-analysis of 17 datasets from Parkinson’s disease GWAS available from European ancestry samples. BP = base-pair coordinates according to human reference genome GRCh38, CHR = Chromosome, dbSNP = database of single nucleotide polymorphisms, L95 = Lower bound of 95% confidence interval for odds ratio, MAF = minor allele frequency, U95= Upper bound of 95% confidence interval for odds ratio.

| CHR | dbSNP ID | BP | MAF |  |  | OR | L95 | U95 | P |
| --- | --- | --- | --- | --- | --- | --- | --- | --- | --- |
|  |  |  | Cases | Controls | All |  |  |  |  |
| 1 | rs114138760 | 154925709 | 0.01 | 0.01 | 0.01 | 1.06 | 0.58 | 1.94 | 0.8446 |
| 1 | rs35749011 | 155162560 | 0.01 | 0.01 | 0.01 | 1.06 | 0.60 | 1.88 | 0.8480 |
| 1 | rs6658353 | 161499264 | 0.49 | 0.47 | 0.48 | 1.00 | 0.88 | 1.13 | 0.9814 |
| 1 | rs11578699 | 171750629 | 0.19 | 0.19 | 0.19 | 1.00 | 0.85 | 1.18 | 0.9792 |
| 1 | rs823118 | 205754444 | 0.42 | 0.42 | 0.42 | 0.97 | 0.85 | 1.10 | 0.5969 |
| 1 | rs11557080 | 205768611 | 0.15 | 0.14 | 0.14 | 1.11 | 0.93 | 1.33 | 0.2516 |
| 1 | rs4653767 | 226728377 | 0.29 | 0.28 | 0.28 | 1.03 | 0.90 | 1.19 | 0.6553 |
| 1 | rs10797576 | 232528865 | 0.14 | 0.14 | 0.14 | 0.94 | 0.79 | 1.13 | 0.5095 |
| 2 | rs76116224 | 17966582 | 0.10 | 0.10 | 0.10 | 1.13 | 0.91 | 1.39 | 0.2654 |
| 2 | rs2042477 | 95335195 | 0.24 | 0.25 | 0.25 | 0.91 | 0.78 | 1.06 | 0.2227 |
| 2 | rs11683001 | 101780501 | 0.31 | 0.31 | 0.31 | 0.90 | 0.78 | 1.03 | 0.1206 |
| 2 | rs57891859 | 134707046 | 0.29 | 0.28 | 0.28 | 1.06 | 0.92 | 1.21 | 0.4523 |
| 2 | rs1474055 | 168253884 | 0.13 | 0.13 | 0.13 | 0.98 | 0.81 | 1.18 | 0.8519 |
| 3 | rs73038319 | 18320267 | 0.04 | 0.05 | 0.05 | 1.02 | 0.74 | 1.41 | 0.9166 |
| 3 | rs6808178 | 28664199 | 0.36 | 0.36 | 0.36 | 1.00 | 0.88 | 1.14 | 0.9947 |
| 3 | rs12497850 | 48711556 | 0.36 | 0.37 | 0.37 | 0.97 | 0.85 | 1.11 | 0.6620 |
| 3 | rs55961674 | 122478045 | 0.17 | 0.17 | 0.17 | 1.01 | 0.85 | 1.20 | 0.9324 |
| 3 | rs11707416 | 151391177 | 0.37 | 0.38 | 0.38 | 0.94 | 0.82 | 1.08 | 0.3755 |
| 3 | rs1450522 | 161359842 | 0.32 | 0.32 | 0.32 | 1.00 | 0.88 | 1.15 | 0.9714 |
| 3 | rs10513789 | 183042285 | 0.20 | 0.21 | 0.21 | 0.95 | 0.82 | 1.12 | 0.5535 |
| 4 | rs873786 | 931588 | 0.10 | 0.11 | 0.11 | 0.90 | 0.73 | 1.11 | 0.3307 |
| 4 | rs34311866 | 958159 | 0.20 | 0.19 | 0.19 | 1.09 | 0.93 | 1.29 | 0.2965 |
| 4 | rs4698412 | 15735725 | 0.45 | 0.46 | 0.46 | 0.95 | 0.84 | 1.08 | 0.4439 |
| 4 | rs34025766 | 17967188 | 0.16 | 0.15 | 0.15 | 0.95 | 0.80 | 1.14 | 0.5962 |
| 4 | rs6825004 | 76189212 | 0.30 | 0.32 | 0.32 | 0.92 | 0.80 | 1.06 | 0.2461 |
| 4 | rs4101061 | 76226816 | 0.28 | 0.29 | 0.29 | 0.96 | 0.84 | 1.11 | 0.6118 |
| 4 | rs6854006 | 76276901 | 0.36 | 0.37 | 0.37 | 0.92 | 0.80 | 1.05 | 0.1909 |
| 4 | rs356182 | 89704960 | 0.33 | 0.34 | 0.34 | 0.93 | 0.81 | 1.07 | 0.3020 |
| 4 | rs5019538 | 89715479 | 0.30 | 0.30 | 0.30 | 0.98 | 0.85 | 1.12 | 0.7275 |
| 4 | rs13117519 | 113447909 | 0.17 | 0.17 | 0.17 | 1.02 | 0.86 | 1.20 | 0.8481 |
| 4 | rs62333164 | 169662006 | 0.33 | 0.34 | 0.34 | 0.96 | 0.84 | 1.10 | 0.5464 |
| 5 | rs1867598 | 60842132 | 0.10 | 0.09 | 0.09 | 1.16 | 0.94 | 1.43 | 0.1691 |
| 5 | rs26431 | 103030090 | 0.29 | 0.30 | 0.30 | 0.95 | 0.83 | 1.10 | 0.5002 |
| 5 | rs11950533 | 134863415 | 0.11 | 0.11 | 0.11 | 1.02 | 0.83 | 1.25 | 0.8480 |
| 6 | rs4140646 | 27771022 | 0.22 | 0.22 | 0.22 | 0.98 | 0.84 | 1.15 | 0.8445 |
| 6 | rs9261484 | 30140906 | 0.24 | 0.26 | 0.25 | 0.95 | 0.82 | 1.10 | 0.4728 |
| 6 | rs504594 | 32610995 | 0.17 | 0.16 | 0.16 | 1.02 | 0.86 | 1.21 | 0.8057 |
| 6 | rs12528068 | 71778059 | 0.28 | 0.29 | 0.28 | 0.96 | 0.84 | 1.11 | 0.6098 |
| 6 | rs997368 | 111922088 | 0.21 | 0.18 | 0.19 | 1.09 | 0.93 | 1.28 | 0.3007 |
| 6 | rs75859381 | 132889222 | 0.02 | 0.03 | 0.03 | 0.92 | 0.61 | 1.39 | 0.6934 |
| 7 | rs199351 | 23260430 | 0.44 | 0.40 | 0.41 | 1.24 | 1.09 | 1.42 | 0.0009 |
| 7 | rs76949143 | 66544864 | 0.05 | 0.04 | 0.05 | 1.14 | 0.85 | 1.52 | 0.3754 |
| 8 | rs1293298 | 11854934 | 0.26 | 0.27 | 0.27 | 0.96 | 0.83 | 1.11 | 0.5876 |
| 8 | rs620513 | 16840084 | 0.29 | 0.28 | 0.28 | 1.02 | 0.88 | 1.17 | 0.8140 |
| 8 | rs2280104 | 22668467 | 0.36 | 0.36 | 0.36 | 1.00 | 0.88 | 1.14 | 0.9926 |
| 8 | rs2086641 | 129889663 | 0.29 | 0.27 | 0.27 | 1.09 | 0.94 | 1.26 | 0.2370 |
| 9 | rs13294100 | 17579692 | 0.35 | 0.36 | 0.36 | 1.01 | 0.88 | 1.15 | 0.9203 |
| 9 | rs10756907 | 17727067 | 0.22 | 0.22 | 0.22 | 0.97 | 0.83 | 1.13 | 0.6608 |
| 9 | rs6476434 | 34046393 | 0.27 | 0.27 | 0.27 | 1.01 | 0.87 | 1.16 | 0.9169 |
| 10 | rs896435 | 15515407 | 0.33 | 0.32 | 0.33 | 1.05 | 0.92 | 1.20 | 0.4620 |
| 10 | rs10748818 | 102255522 | 0.15 | 0.15 | 0.15 | 1.04 | 0.88 | 1.24 | 0.6374 |
| 10 | rs72840788 | 119656173 | 0.20 | 0.21 | 0.21 | 1.01 | 0.87 | 1.18 | 0.8767 |
| 10 | rs117896735 | 119776815 | 0.01 | 0.02 | 0.02 | 0.69 | 0.40 | 1.21 | 0.1930 |
| 11 | rs7938782 | 10537230 | 0.14 | 0.12 | 0.12 | 1.12 | 0.93 | 1.34 | 0.2509 |
| 11 | rs12283611 | 83776234 | 0.44 | 0.42 | 0.42 | 1.08 | 0.95 | 1.23 | 0.2307 |
| 11 | rs3802920 | 133917106 | 0.20 | 0.19 | 0.19 | 1.01 | 0.86 | 1.18 | 0.9229 |
| 12 | rs76904798 | 40220632 | 0.15 | 0.14 | 0.14 | 1.13 | 0.95 | 1.35 | 0.1767 |
| 12 | rs7134559 | 46025303 | 0.41 | 0.40 | 0.40 | 0.98 | 0.86 | 1.12 | 0.8099 |
| 12 | rs10847864 | 122842051 | 0.34 | 0.34 | 0.34 | 0.97 | 0.85 | 1.11 | 0.6768 |
| 12 | rs11610045 | 132487182 | 0.48 | 0.50 | 0.50 | 0.94 | 0.82 | 1.07 | 0.3192 |
| 13 | rs9568188 | 49353596 | 0.26 | 0.27 | 0.27 | 1.01 | 0.87 | 1.16 | 0.9488 |
| 13 | rs4771268 | 97212767 | 0.23 | 0.22 | 0.22 | 1.02 | 0.88 | 1.19 | 0.7802 |
| 14 | rs12147950 | 37520065 | 0.44 | 0.44 | 0.44 | 0.95 | 0.83 | 1.08 | 0.4011 |
| 14 | rs11158026 | 54882151 | 0.35 | 0.32 | 0.32 | 1.13 | 0.98 | 1.29 | 0.0841 |
| 14 | rs3742785 | 74906331 | 0.22 | 0.21 | 0.21 | 1.06 | 0.91 | 1.24 | 0.4377 |
| 14 | rs979812 | 87997920 | 0.47 | 0.43 | 0.43 | 1.17 | 1.03 | 1.33 | 0.0195 |
| 15 | rs2251086 | 61705186 | 0.15 | 0.16 | 0.16 | 1.01 | 0.84 | 1.20 | 0.9557 |
| 16 | rs6497339 | 19266171 | 0.44 | 0.46 | 0.46 | 0.93 | 0.82 | 1.05 | 0.2523 |
| 16 | rs2904880 | 28933075 | 0.32 | 0.33 | 0.32 | 1.00 | 0.87 | 1.15 | 0.9944 |
| 16 | rs11150601 | 30966478 | 0.38 | 0.36 | 0.37 | 1.09 | 0.95 | 1.24 | 0.2190 |
| 16 | rs6500328 | 50702745 | 0.41 | 0.40 | 0.40 | 1.02 | 0.89 | 1.16 | 0.8151 |
| 16 | rs3104783 | 52602330 | 0.44 | 0.42 | 0.42 | 1.07 | 0.94 | 1.21 | 0.3111 |
| 16 | rs10221156 | 52935514 | 0.10 | 0.10 | 0.10 | 0.97 | 0.78 | 1.20 | 0.7499 |
| 17 | rs12600861 | 7452302 | 0.36 | 0.35 | 0.35 | 1.02 | 0.89 | 1.17 | 0.7553 |
| 17 | rs12951632 | 42588995 | 0.28 | 0.26 | 0.27 | 1.07 | 0.93 | 1.24 | 0.3285 |
| 17 | rs2269906 | 44216969 | 0.35 | 0.33 | 0.33 | 1.10 | 0.96 | 1.25 | 0.1761 |
| 17 | rs850738 | 44357262 | 0.39 | 0.42 | 0.41 | 0.90 | 0.79 | 1.03 | 0.1150 |
| 17 | rs62053943 | 45666837 | 0.17 | 0.15 | 0.15 | 1.09 | 0.92 | 1.29 | 0.3456 |
| 17 | rs117615688 | 45720942 | 0.07 | 0.06 | 0.06 | 0.99 | 0.78 | 1.27 | 0.9590 |
| 17 | rs11658976 | 46789439 | 0.42 | 0.40 | 0.40 | 1.13 | 0.99 | 1.29 | 0.0635 |
| 17 | rs61169879 | 61840005 | 0.18 | 0.16 | 0.17 | 1.18 | 1.00 | 1.40 | 0.0458 |
| 17 | rs666463 | 78429399 | 0.14 | 0.17 | 0.16 | 0.76 | 0.63 | 0.91 | 0.0026 |
| 18 | rs1941685 | 33724354 | 0.52 | 0.49 | 0.49 | 1.13 | 0.99 | 1.28 | 0.0610 |
| 18 | rs12456492 | 43093415 | 0.32 | 0.32 | 0.32 | 1.02 | 0.89 | 1.17 | 0.7305 |
| 18 | rs8087969 | 51157219 | 0.46 | 0.46 | 0.46 | 1.03 | 0.91 | 1.17 | 0.5991 |
| 19 | rs55818311 | 2341049 | 0.32 | 0.32 | 0.32 | 0.99 | 0.87 | 1.14 | 0.9333 |
| 20 | rs77351827 | 6025395 | 0.15 | 0.12 | 0.13 | 1.21 | 1.01 | 1.45 | 0.0397 |
| 21 | rs2248244 | 37480059 | 0.26 | 0.27 | 0.27 | 0.97 | 0.84 | 1.12 | 0.7098 |

## Supplementary Figures

**Supplementary Fig. S1.**
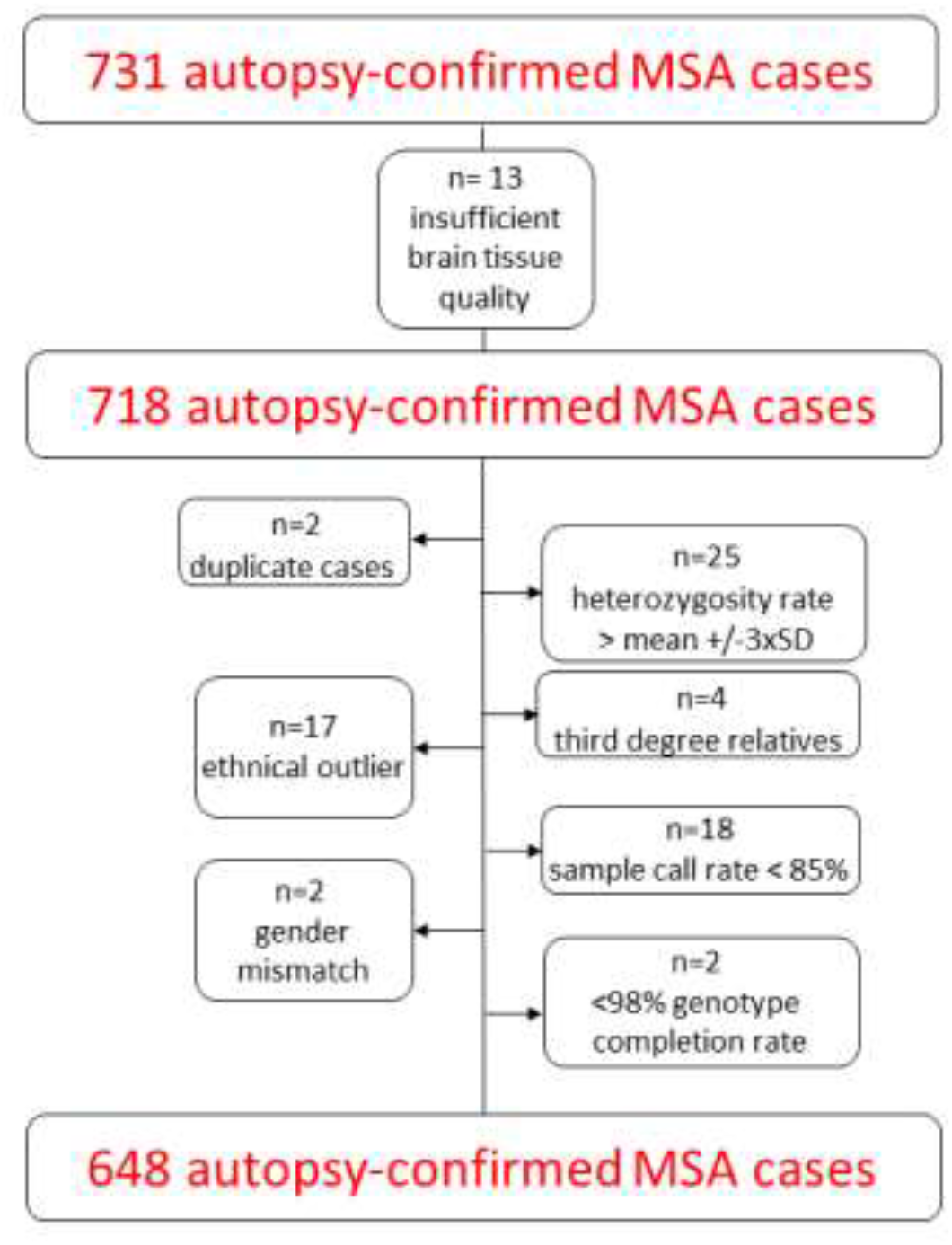
Flowchart sample quality control. SD = standard deviation.

**Supplementary Fig. S2.**
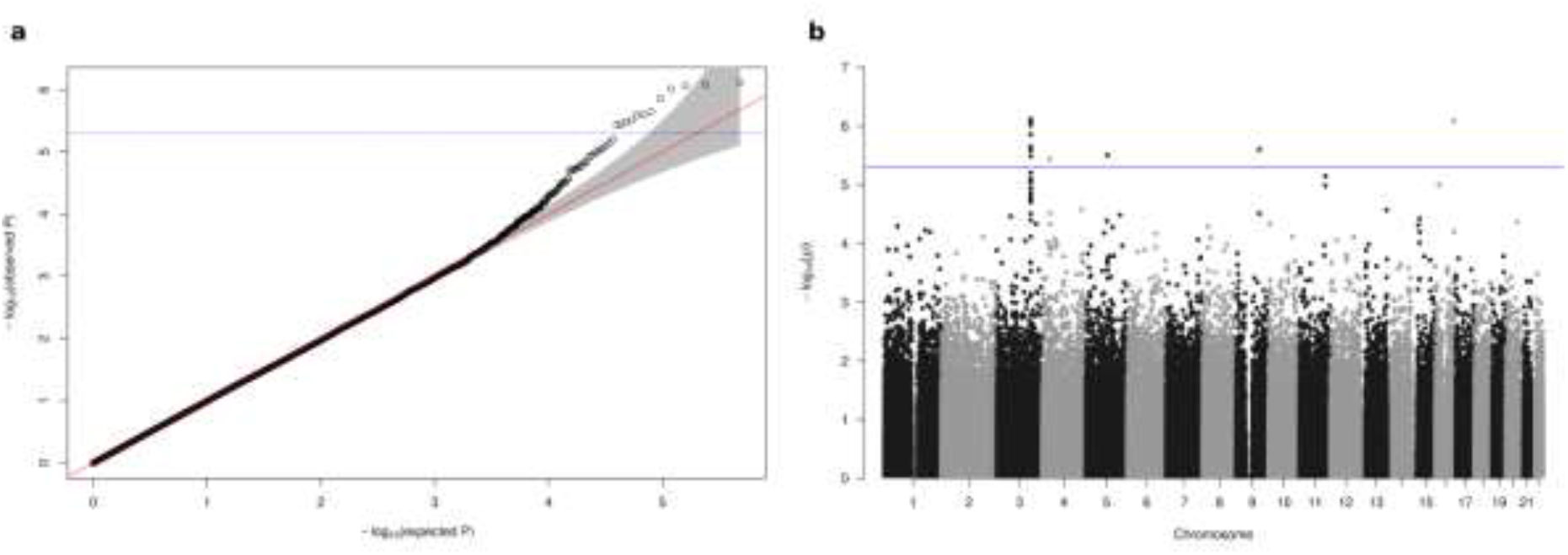
Association plots for genotyped data. **(a)** QQ plot based on 477,261 variants. (**b)** Manhattan plot showing –log10 P-values from logistic regression on genotyped variants with sex and two principal components as covariates plotted against their chromosomal position.

**Supplementary Fig. S3.**
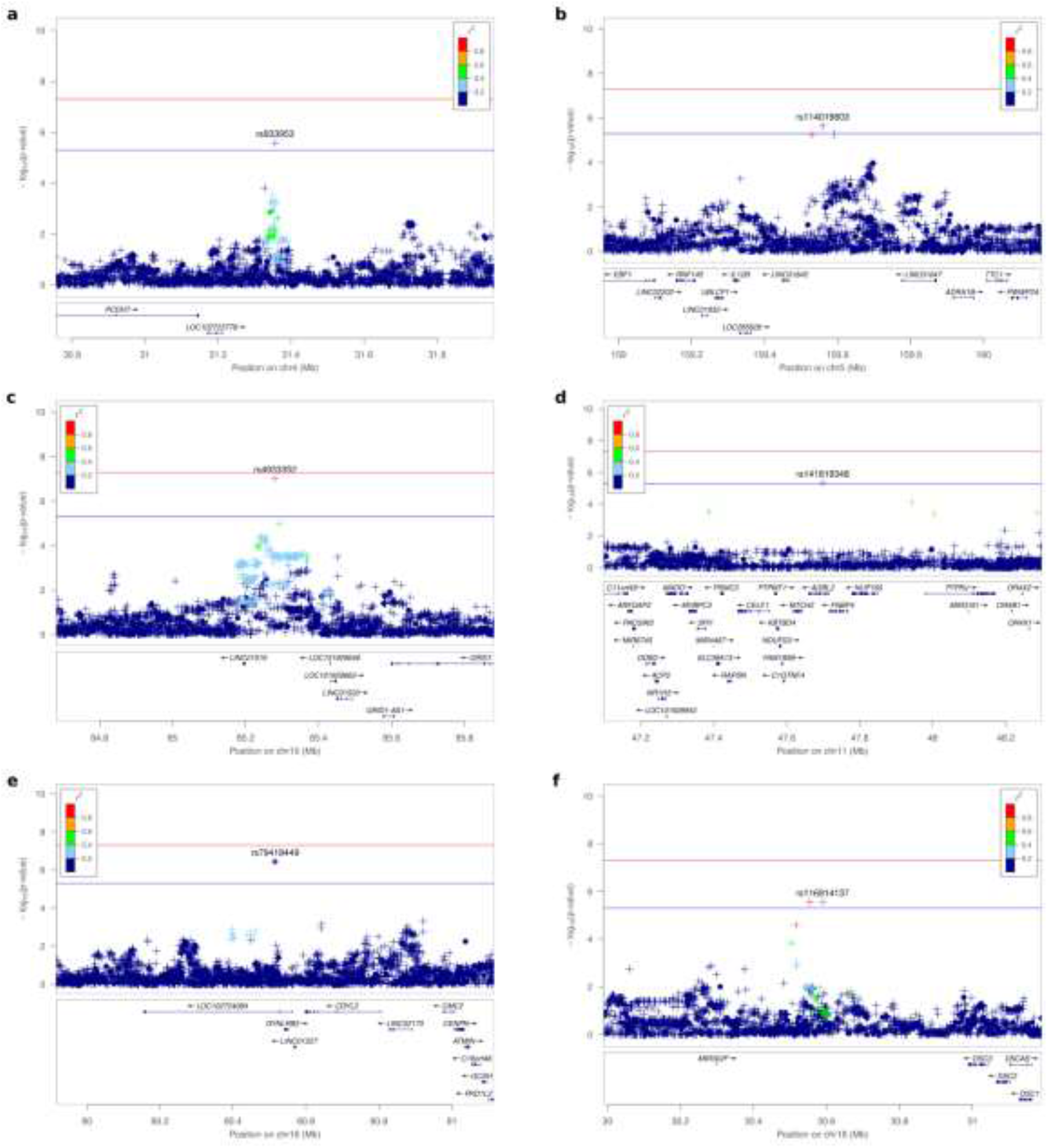
Regional plots for further loci with suggestive associations. **(a)** Regional plot for associations on chromsome 4 in the genomic region from 30.8 to 32.0 Mb. **(b)** Regional plot for associations on chromsome 5 in the genomic region from 159.0 to 160.2 Mb. **(c)** Regional plot for associations on chromsome 10 in the genomic region from 84.7 to 85.9 Mb. **(d)** Regional plot for associations on chromsome 11 in the genomic region from 47.1 to 48.3 Mb. **(e)** Regional plot for associations on chromsome 16 in the genomic region from 79.9 to 81.1 Mb. **(f)** Regional plot for associations on chromsome 18 in the genomic region from 30.0 to 31.2 Mb.

